# Pretend play predicts receptive and expressive language trajectories in young children with autism

**DOI:** 10.1101/2022.04.04.22273397

**Authors:** Andrey Vyshedskiy, Edward Khokhlovich

## Abstract

The effect of pretend play in 2 to 5-year-old children with ASD was investigated in the largest and the longest observational study to-date. Parents assessed the development of 7,069 children quarterly for three years on five subscales: combinatorial receptive language, expressive language, sociability, sensory awareness, and health. Pretend play was associated with superior developmental trajectories: 1.9-fold faster improvement of combinatorial receptive language (p<0.0001), 1.4-fold faster improvement of expressive language (p<0.0001), and 1.3-fold faster improvement of sensory awareness (p=0.0009). Pretend play had little effect on sociability and health. The strong association of pretend play with combinatorial receptive language remained significant even when controlling for expressive language. Similarly, the effect of pretend play on expressive language remained significant even when controlling for combinatorial receptive language. The effect of pretend play on combinatorial receptive language (but not on the expressive language) was stronger than the effects of seizures, sleep problems or high-TV exposure. The strong effect of pretend-play supports earlier studies indicating that it is an important stepping stone for language acquisition, particularly, the acquisition of combinatorial language.

## Introduction

Pretend play is separated from other forms of daily activities by its creative aspect, its difference from habitual actions and those performed for the sake of necessity. Pretend play is exhibited by children universally across all cultures and begins unprompted between the 1.5 to 2 years of age (Lillard et al., 2011). The absence of pretend play puts a child at high risk for ASD and represents a key deficit used in the diagnosis of ASD (Baron-Cohen et al., 1992; Osterling & Dawson, 1994). Pretend play has been also theorized to have a predictive relationship with expressive and receptive language development in children (Lewis, 2000; Stagnitti & Lewis, 2015; Toth et al., 2006) and there is a significant interest in symbolic play interventions (Kasari et al., 2006).

Most studies investigating the effect of pretend play, however, suffer from low numbers of participants. In 2015 we published a language training app for children (Dunn, Elgart, Lokshina, Faisman, Khokhlovich, et al., 2017b, 2017a; Dunn, Elgart, Lokshina, Faisman, Waslick, et al., 2017; Vyshedskiy, Khokhlovich, et al., 2020; Vyshedskiy & Dunn, 2015a), which invites parents to complete their child’s evaluation every three months. As a result, we accumulated over hundred thousand comprehensive longitudinal evaluations on five subscales (combinatorial receptive language, expressive language, sociability, sensory awareness, and health) that give us an opportunity to study children’s developmental trajectories. In this report, we investigate the effect of pretend play reported by parents in response to the question: “My child engages in a VARIETY of make-believe activities (such as: playing house, playing with toy soldiers, building forts and castles, etc.).” The possible answers were: “very true,” “somewhat true,” and “not true.” To assess the effect of pretend play, we compared participants who answered “very true” to participants who answered “not true” over all available subscales: receptive language, expressive language, sociability, sensory awareness, and health.

Expressive language, sociability, sensory awareness, and health were assessed by Autism Treatment Evaluation Checklist (ATEC) (Rimland & Edelson, 1999), a measure validated for longitudinal tracking of symptoms and assessing changes in ASD severity (Charman et al., 2004; Klaveness et al., 2013; Magiati et al., 2011; Mahapatra, Khokhlovich, et al., 2018). The expressive language subscale of ATEC consists of 14 items; the sociability subscale contains 20 items; the sensory awareness subscale has 18 items; and the health subscale contains 25 items.

Receptive language was assessed with Mental Synthesis Evaluation Checklist (MSEC) (Braverman et al., 2018), a 20-item questionnaire that does not measure knowledge of individual words but focuses on combinatorial language skills. MSEC items include questions like: Understands simple stories that are read aloud; Understands elaborate fairy tales that are read aloud; Understands some simple modifiers; Understands several modifiers in a sentence; Understands size; Understands spatial prepositions; Understands the change in meaning when the order of words is changed; Understands explanations about people, objects or situations beyond the immediate surroundings; and so on. MSEC has been shown to have good internal reliability, adequate test–retest reliability, good construct validity, and good known group validity (Braverman et al., 2018).

The framework for the evaluation of score changes over time was developed in Mahapatra *et al*. (Mahapatra, Khokhlovich, et al., 2018) and further validated in Vyshedskiy *et al*. (Vyshedskiy, Khokhlovich, et al., 2020) and Fridberg *et al*.(Fridberg et al., 2021). This robust technique uses the Linear Mixed Effect Model with Repeated Measures to model complex relationships between time of evaluation, group assignment (Pretend Play or no Pretend Play), and evaluation subscales. The model then computes averages for each group and for each subscale at each time point.

## Methods

### Participants

Participants were users of a language therapy app that was made available gratis at all major app stores in September 2015. Once the app was downloaded, caregivers were asked to register and to provide demographic details, including the child’s diagnosis and age. Caregivers consented to anonymized data analysis and completed the Autism Treatment Evaluation Checklist (ATEC) (Rimland & Edelson, 1999), an evaluation of the receptive language using the Mental Synthesis Evaluation Checklist (MSEC) (Braverman et al., 2018), as well as the Screen Time assessment and the Diet and Supplements assessment. The first evaluation was administered approximately one month after the download. The subsequent evaluations were administered at approximately three-month intervals. To enforce regular evaluations, the app became unusable at the end of each three-month interval and parents were required to complete an evaluation to regain its functionality.

#### Inclusion criteria

Inclusion criteria were identical to our previous studies of this population (Fridberg et al., 2021; Mahapatra, Khokhlovich, et al., 2018; Vyshedskiy, Khokhlovich, et al., 2020). Specifically, we selected participants based on the following criteria:

1. Consistency: Participants must have filled out at least three ATEC evaluations and the interval between the first and the last evaluation was six months or longer.
2. Diagnosis: ASD. Children without ASD diagnosis were excluded from the study. Other diagnostic options included: Mild Language Delay, Pervasive Developmental Disorder, Attention Deficit Disorder, Social Communication Disorder, Specific Language Impairment, Apraxia, Sensory Processing Disorder, Down Syndrome, Lost Diagnosis of Autism or PDD, Other Genetic Disorder, normally developed child. A good reliability of parent-reported diagnosis was previously demonstrated in Ref. (Jagadeesan et al., n.d.).The parent-reported ASD diagnosis was also supported by ATEC scores. Average initial ATEC total score was 68.5 ± 25.1, which corresponds to moderate-to-severe ASD as delineated in Ref. (Mahapatra, Vyshedsky, et al., 2018) and Table 1.

**Table 1.** **Approximate relationship between ATEC total score, age, and ASD severity as described in Mahapatra *et al***. (Mahapatra, Khokhlovich, et al., 2018). **At any age, a greater ATEC score indicates greater ASD severity.**

| Severity | Age |  |  |  |  |  |  |  |  |  |  |
| --- | --- | --- | --- | --- | --- | --- | --- | --- | --- | --- | --- |
|  | 2 | 3 | 4 | 5 | 6 | 7 | 8 | 9 | 10 | 11 | 12 |
| **Mild** | <82 | <65 | <52 | <43 | <36 | <31 | <28 | <25 | <23 | <21 | <20 |
| **Moderate** | 82-130 | 65-103 | 52-83 | 43-69 | 36-58 | 31-50 | 28-44 | 25-39 | 23-36 | 21-34 | 20-32 |
| **Severe** | 130-179 | 103-179 | 83-179 | 69-179 | 58-179 | 50-179 | 44-179 | 39-179 | 36-179 | 34-179 | 32-179 |

#### Exclusion criteria

1. Maximum age: Participants older than five years of age at the time of their first evaluation were excluded from this study.
2. Minimum age: Participants who completed their first evaluation before the age of two years were excluded from this study.

After excluding participants that did not meet these criteria, there were 7,069 total participants.

### Pretend play assessment

Parents were required to respond to the question: “[My child] Engages in a VARIETY of make-believe activities (such as: playing house, playing with toy soldiers, building forts and castles, etc.).” The possible answers were: “very true,” “somewhat true,” and “not true.” To assess the effect of pretend play, we compared participants who answered “very true” (N=2258; the Pretend Play group, abbreviated PP) to participants who answered “not true” (N=4811; the no Pretend Play group, abbreviated noPP).

### Evaluations

A caregiver-completed Autism Treatment Evaluation Checklist (ATEC) (Rimland & Edelson, 1999) and Mental Synthesis Evaluation Checklist (MSEC) (Braverman et al., 2018) were used to track child development. The ATEC questionnaire is comprised of four subscales: 1) Speech/Language/Communication, 2) Sociability, 3) Sensory/Cognitive Awareness, and 4) Physical/Health/Behavior. The first subscale, Speech/Language/Communication, contains 14 items and its score ranges from 0 to 28 points. The Sociability subscale contains 20 items within a score range of 0 to 40 points. The third subscale, referred here as the Sensory Awareness subscale, has 18 items and score range from 0 to 36 points. The fourth subscale, referred here as the Health subscale contains 25 items and score range from 0 to 75 points. The scores from each subscale are combined in order to calculate a Total Score, which ranges from 0 to 179 points. A lower score indicates lower severity of ASD symptoms and a higher score indicates more severe symptoms of ASD. ATEC is not a diagnostic checklist. It was designed to evaluate the effectiveness of treatment (Rimland & Edelson, 1999), thus ASD severity can only have an approximate relationship with the total ATEC score and age. Table 1 lists approximate ATEC total score as related to ASD severity and age as described in Mahapatra *et al*. (Mahapatra, Khokhlovich, et al., 2018).

ATEC was selected because it is one of the few measures validated to evaluate treatment effectiveness. In contrast, another popular ASD assessment tool, Autism Diagnostic Observation Schedule or ADOS, (Lord et al., 2000) has only been validated as a diagnostic tool. Various studies confirmed the validity and reliability of ATEC (Al Backer, 2016; Geier et al., 2013; Jarusiewicz, 2002) and several trials confirmed ATEC’s ability to longitudinally measure changes in participant performance (Charman et al., 2004; Klaveness et al., 2013; Magiati et al., 2011; Mahapatra, Khokhlovich, et al., 2018). Whitehouse et al. used ATEC as a primary outcome measure for a randomized controlled trial of their iPad-based intervention for ASD, named “Therapy Outcomes By You” or TOBY, and noted ATEC’s “internal consistency and adequate predictive validity” (Whitehouse et al., 2017). These studies support the effectiveness of ATEC as a tool for longitudinal tracking of symptoms and assessing changes in ASD severity.

### Expressive language assessment

The ATEC Speech/Language/Communication subscale includes the following questions: 1) Knows own name, 2) Responds to ‘No’ or ‘Stop’, 3) Can follow some commands, 4) Can use one word at a time (No!, Eat, Water, etc.), 5) Can use 2 words at a time (Don’t want, Go home), 6) Can use 3 words at a time (Want more milk), 7) Knows 10 or more words, 8) Can use sentences with 4 or more words, 9) Explains what he/she wants, 10) Asks meaningful questions, 11) Speech tends to be meaningful/relevant, 12) Often uses several successive sentences, 13) Carries on fairly good conversation, and 14) Has normal ability to communicate for his/her age. With the exception of the first three items, all items in the ATEC subscale 1 primarily measure expressive language. Accordingly, the ATEC subscale 1 is herein referred to as the Expressive Language subscale to distinguish it from the Receptive Language subscale tested by the MSEC evaluation.

### Receptive language assessment

The MSEC evaluation was designed to be complementary to ATEC in measuring combinatorial receptive language. Out of 20 MSEC items, those that directly assess receptive language are the following: 1) Understands simple stories that are read aloud; 2) Understands elaborate fairy tales that are read aloud (i.e., stories describing FANTASY creatures); 3) Understands some simple modifiers (i.e., green apple vs. red apple or big apple vs. small apple); 4) Understands several modifiers in a sentence (i.e., small green apple); 5) Understands size (can select the largest/smallest object out of a collection of objects); 6) Understands possessive pronouns (i.e. your apple vs. her apple); 7) Understands spatial prepositions (i.e., put the apple ON TOP of the box vs. INSIDE the box vs. BEHIND the box); 11) Understands verb tenses (i.e., I will eat an apple vs. I ate an apple); 8) Understands the change in meaning when the order of words is changed (i.e., understands the difference between ‘a cat ate a mouse’ vs. ‘a mouse ate a cat’); 9) Understands explanations about people, objects or situations beyond the immediate surroundings (e.g., “Mom is walking the dog,” “The snow has turned to water”). MSEC consists of 20 questions within a score range of 0 to 40 points; similarly to ATEC, a lower MSEC score indicates a better developed receptive language.

The psychometric quality of MSEC was tested with 3,715 parents of ASD children (Braverman et al., 2018). Internal reliability of MSEC was good (Cronbach’s alpha > 0.9). MSEC exhibited adequate test–retest reliability, good construct validity, and good known group validity reflected by the difference in MSEC scores for children of different ASD severity levels.

To simplify interpretation of figure labels, the subscale 1 of the ATEC evaluation is herein referred to as the Expressive Language subscale and the MSEC scale is referred to as the Receptive Language subscale.

### Statistical analysis

The framework for the evaluation of score changes over time was explained in detail in Mahapatra *et al*. (Mahapatra, Khokhlovich, et al., 2018) and Vyshedskiy *et al*. (Vyshedskiy, Khokhlovich, et al., 2020). In short, the concept of a “Visit” was developed by dividing the three-year-long observation interval into 3-month periods. All evaluations were mapped into 3-month-long bins with the first evaluation placed in the first bin. When more than one evaluation was completed within a bin, their results were averaged to calculate a single number representing this 3-month interval. Thus, we had 12 quarterly evaluations for both PP and noPP groups.

It was then hypothesized that there was a two-way interaction between pretend-play-group and Visit. Statistically, this hypothesis was modeled by applying the Linear Mixed Effect Model with Repeated Measures (MMRM), where a two-way interaction term was introduced to test the hypothesis. The model (Endpoint ∼ Baseline + Gender + Severity + Pretend-Play-Group * Visit) was fit using the R Bioconductor library of statistical packages, specifically the “nlme” package. The subscale score at baseline, as well as gender and severity were used as covariates. Conceptually, the model fits a plane into n-dimensional space. This plane considers a complex variability structure across multiple visits, including baseline differences. Once such a plane is fit, the model calculates Least Squares Means (LS Means) for each subscale and group at each visit. The model also calculates LS Mean differences between the groups at each visit.

In preparation for statistical analysis, participants in the PP group were matched to those in the noPP group using propensity score (Schneider et al., 2007) based on age, gender, expressive language, receptive language, sociability, sensory awareness, and health at the 1^st^ evaluation (baseline). The number of matched participants was 2,254 in each group, Table 2.

**Table 2.** Demographic data of participant pool.

|  | **Pretend play group** | **No pretend play** |
| --- | --- | --- |
| **Number of participants** | 2,258 | 4,811 |
| **Number of matched participants** | 2,254 | 2,254 |
| **Age at baseline** | 3.4±0.8 | 3.4±0.8 |
| **Male Gender** | 68% | 83% |

### Community involvement statement

Autistic people and family members were involved in the design of the combinatorial receptive language outcome measure MSEC.

## Results

On the Receptive Language subscale, the average improvement in the Pretend Play (PP) group over 36 months was 11.31 points (SE=0.55, p<0.0001) compared to 5.88 points (SE=0.48, p<0.0001) in the no Pretend Play (noPP) group, Figure 1A, Table 3, Table S1. The difference in the PP group relative to the noPP group at Month 36 was statistically significant: -7.26 points (SE=0.72, p<0.0001). The negative difference (marked in the Table 3 as “PP – noPP”) indicates that the noPP group had greater scores at Month 36 and, therefore, more severe symptoms. On an annualized basis, the participants from PP group improved their receptive language 1.9-times faster than those in the noPP group (PP group =3.8 points/year; noPP group =2.0 points/year).

**Table 3.** **Characteristics of Pretend Play (PP) and no Pretend Play groups (noPP). Data is presented as LS Means (SE; 95% CI). A lower score indicates a lower severity of ASD symptoms. The difference between Pretend Play and No Pretend Play (PP – noPP) is presented as: LS Mean (SE; P-value). The negative PP – noPP difference indicates that noPP group had higher score and therefore more severe symptoms**.

| Subscale | Baseline |  |  | Month 36 |  |  | Month 36 – Baseline |  |
| --- | --- | --- | --- | --- | --- | --- | --- | --- |
|  | Pretend Play | No Pretend Play | PP – NoPP | Pretend Play | No Pretend Play | PP – NoPP | Pretend Play | No Pretend Play |
| **Receptive Language** | 27<br>(0.134;<br>26.7 -<br>27.2) | 28.8 (0.13;<br>28.5 - 29.1) | -1.83<br>(0.18;<br><0.0001) | 15.7<br>(0.546;<br>14.6 -<br>16.7) | 22.9<br>(0.474; 22 -<br>23.8) | -7.26<br>(0.72;<br><0.0001) | -11.31<br>(0.55;<br><0.0001) | -5.88 (0.48;<br><0.0001) |
| **Expressive Language** | 14.98<br>(0.0993;<br>14.78 -<br>15.17) | 15.77<br>(0.0971;<br>15.58 -<br>15.96) | -0.79<br>(0.13;<br><0.0001) | 7.3<br>(0.4018;<br>6.51 -<br>8.09) | 10.25<br>(0.3488;<br>9.57 -<br>10.94) | -2.95<br>(0.53;<br><0.0001) | -7.68 (0.4;<br><0.0001) | -5.52 (0.35;<br><0.0001) |
| **Sociability** | 13.5<br>(0.127;<br>13.2 -<br>13.7) | 13.4 (0.124;<br>13.1 - 13.6) | 0.13<br>(0.17;<br>0.454) | 10.9<br>(0.541;<br>9.8 -<br>11.9) | 11.1 (0.47;<br>10.2 - 12) | -0.24<br>(0.72;<br>0.7348) | -2.63<br>(0.55;<br><0.0001) | -2.26 (0.48;<br><0.0001) |
| **Sensory Awareness** | 12.77<br>(0.104;<br>12.56 -<br>12.97) | 14.15<br>(0.103;<br>13.95 -<br>14.35) | -1.39<br>(0.14;<br><0.0001) | 10.35<br>(0.453;<br>9.46 -<br>11.24) | 12.34<br>(0.393;<br>11.57 -<br>13.11) | -1.99 (0.6;<br>0.0009) | -2.41<br>(0.46;<br><0.0001) | -1.81 (0.4;<br><0.0001) |
| **Health** | 21.4<br>(0.193;<br>21 - 21.8) | 20.3 (0.187;<br>19.9 - 20.6) | 1.11<br>(0.25;<br><0.0001) | 20.0<br>(0.815;<br>18.5 -<br>21.6) | 19.3<br>(0.707;<br>17.9 - 20.7) | 0.76 (1.08;<br>0.4782) | -1.33<br>(0.82;<br>0.1062) | -0.98 (0.72;<br>0.1699) |

**Figure 1.**
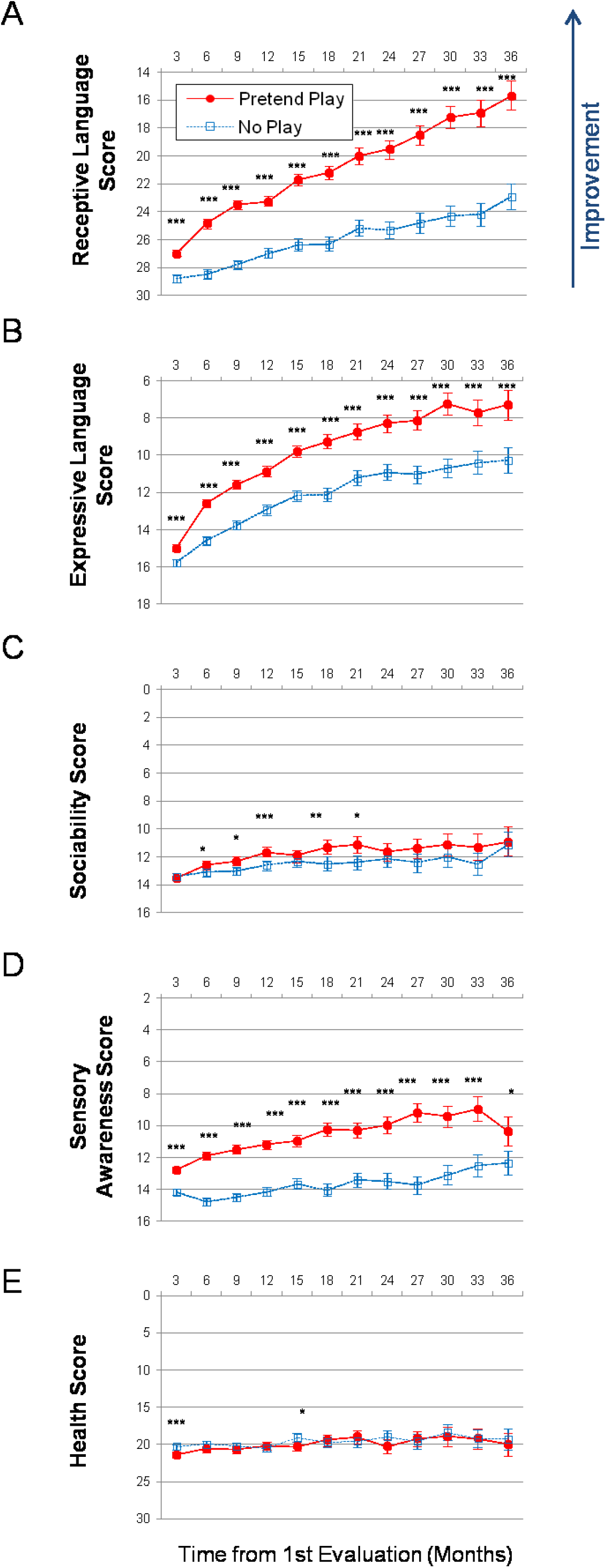
**Longitudinal plots of subscale scores LS Means. Horizontal axis shows months from the 1st evaluation (0 to 36 months). Error bars show the 95% confidence interval. To facilitate comparison between subscales, all vertical axes ranges have been normalized to show 40% of their corresponding subscale’s maximum available score. A lower score indicates symptom improvement. P-value is marked: ***<0.0001; **<0.001; *<0.05. (A) Receptive Language score. (B) Expressive Language score. (C) Sociability score. (D) Sensory Awareness score (E) Health score.**

On the Expressive Language subscale, the participants in the PP group improved over the 36-month period by 7.68 points (SE=0.4, p<0.0001) compared to 5.52 points (SE=0.35, p<0.0001) improvement in the noPP group, Figure 1B, Table S2. The difference in the PP group relative to the noPP group at Month 36 was: -2.95 points (SE=0.53, p<0.0001). On an annualized basis, the subjects in the PP group improved their expressive language 1.4-times faster than those in the noPP group (PP group =2.6 points/year; noPP group =1.8 points/year).

On the Sociability subscale, the subjects in the PP group improved over the 36-month period by 2.63 points (SE=0.55, p<0.0001) compared to 2.26 points (SE=0.48, p<0.0001) improvement in those in the noPP group, Figure 1C, Table S3. The difference in the PP group relative to the noPP group at Month 36 was not statistically significant: -0.24 points (SE=0.72, p=0.7348).

On the Sensory Awareness subscale, the subjects in the PP group improved over the 36-month period by 2.41 points (SE=0.46, p<0.0001) compared to 1.81 points (SE=0.4, p<0.0001) improvement in the noPP group, Figure 1D, Table S4. The difference in the PP group relative to the noPP group at Month 36 was: -1.99 points (SE=0.6, p=0.0009). On an annualized basis, the participants in the PP group improved their sensory awareness 1.3-times faster than those in the PP group (PP group =0.8 points/year; noPP group =0.6 points/year).

On the Health subscale, the subjects in the PP group improved over the 36-month period by 1.33 points (SE=0.82, p=0.1062) compared to 0.98 points (SE=0.72, p=0.1699) improvement in the noPP group, Figure 1E, Table S5. The difference in the PP group relative to the noPP group at Month 36 was not statistically significant: 0.76 points (SE=1.08, p=0.4782).

The strong association of pretend play with receptive language remained significant even when controlling for expressive language (in addition to the receptive language score at baseline as well as gender and severity). The average improvement in the subjects in the PP group over 36 months was 8.42 points (SE=0.53, p<0.0001) compared to 3.69 points (SE=0.46, p<0.0001) in the noPP group, Table S6. The difference in the PP group relative to the noPP group at Month 36 was statistically significant: -6.09 points (SE=0.69, p<0.0001). On an annualized basis, the participants in the PP group improved their receptive language 2.3-times faster than those in the noPP group, when controlling for expressive language (PP group =2.8 points/year; NoPP group =1.2 points/year).

Similarly, the effect of pretend play on expressive language remained significant even when controlling for receptive language (in addition to the expressive language score at baseline as well as gender and severity). The subjects in the PP group improved over the 36-month period by 5.39 points (SE=0.39, p<0.0001) compared to 4.38 points (SE=0.34, p<0.0001) improvement in the noPP group, Table S7. The difference in the PP group relative to noPP group at Month 36 was: -1.15 points (SE=0.51, p<0.0253). On an annualized basis, the participants in the PP group improved their expressive language 1.2-times faster than those in the noPP group (PP group =1.8 points/year; noPP group =1.5 points/year).

To put the effect of pretend play into perspective, we have compared its magnitude with the effects of seizures (Forman et al., 2022), sleep problems (Levin et al., 2022), and high TV exposure (Fridberg et al., 2021) at Month 36, Table 4. The Receptive Language score difference in the PP group relative to noPP group at Month 36 was significantly greater (−7.26 points, SE=0.72, p<0.0001) compared to the effect of seizures (−0.43 points, i.e., 17 times smaller than the effect of PP; SE=1.38; p=0.7537), effect of sleep problems (−0.9 points, i.e., 8 times smaller than the effect of PP; SE=1.09; p=0.406), and the effect of high-TV-exposure (−2.58 points, i.e., 3 times smaller than the effect of PP; SE=1.04; p=0.0128). The Expressive Language score difference in the PP group relative to noPP group at Month 36 was similar (−2.95 points, SE=0.53, p<0.0001) compared to the effect of seizures (−2.61 points, SE=0.9; p=0.0037), the effect of sleep problems (−0.12 points, SE=0.77; p=0.8728), and the effect of high-TV-exposure (−1.26 points, SE=0.7; p=0.0719).

**Table 4.** **The effect of pretend play is compared to the effect of other health conditions at Month 36. The difference between groups is presented as: LS Mean (SE; P-value). The negative difference indicates a lower score (and therefore greater improvement) in the 1**^**st**^ **group (PP) compared to the 2**^**nd**^ **group (noPP). Participants with seizures (mild, moderate or severe seizures as reported by parents, N=971) were matched to those with no seizures based on age, gender, expressive language, receptive language, sociability, sensory awareness, and health at the 1**^**st**^ **evaluation (baseline)** (Forman et al., 2022). **Participants with sleep problems (moderate or severe sleep problems as reported by parents; N=1030) were matched to those with no sleep problems based on age, gender, expressive language, receptive language, sociability, sensory awareness, and health at the 1**^**st**^ **evaluation** (Levin et al., 2022). **High-TV users (2 hours or more of videos and television per day; N=797) were matched to low-TV users (40 minutes or less of videos and television per day) based on age, gender, expressive language, receptive language, sociability, sensory awareness, and health at the 1**^**st**^ **evaluation** (Fridberg et al., 2021).

| **Month 36** | **Pretend Play – No Pretend Play** | **No Seizures – Seizures** | **No Sleep Problems – Sleep Problems** | **Low-TV use – High-TV use** |
| --- | --- | --- | --- | --- |
| **Receptive Language** | -7.26 (0.72; <0.0001) | -0.43 (1.38; 0.7537) | -1.63 (1.28; 0.2022) | -2.58 (1.04; 0.0128) |
| **Expressive Language** | -2.95 (0.53; <0.0001) | -2.61 (0.9; 0.0037) | -1.62 (0.94; 0.0856) | 1.26 (0.7; 0.0719) |
| **Sociability** | -0.24 (0.72; 0.7348) | -2.58 (1.2; 0.032) | -2.67 (1.31; 0.0426) | -1.82 (0.99; 0.0663) |
| **Sensory Awareness** | -1.99 (0.6; 0.0009) | -2.97 (1.05; 0.0047) | -1.51 (1.11; 0.1751) | -1.58 (0.85; 0.0631) |
| **Health** | 0.76 (1.08; 0.4782) | -8.29 (2.02; <0.0001) | -14.14 (1.93; <0.0001) | -1.05 (1.52; 0.4898) |

## Discussion

This is the largest study to date demonstrating a strong association between pretend play and language. By Month 36, the participants in pretend play (PP) group outperformed those in no pretend play (noPP) group by a factor of 1.9 in combinatorial receptive language (7.26 points, SE=0.72; P-value<0.0001); by a factor of 1.4 in expressive language (2.95 points, SE=0.53; P-value<0.0001); and by a factor of 1.3 in sensory awareness (1.99 points, SE=0.6; P-value=0.0009), Figure 1, Table 3. The difference in sociability and health was not statistically significant.

How does 1.9-fold difference in combinatorial receptive language acquisition compare to the effect of other health ailments? We have previously investigated the effects of seizures (Forman et al., 2022), sleep problems (Levin et al., 2022), and high TV exposure (Fridberg et al., 2021) on a child’s developmental trajectory. Accordingly we were able to compare the receptive language advantage associated with pretend play to the effect of seizures, sleep problems, and high TV exposure (Table 4). To our surprise, in terms of the absolute value, the positive effect of pretend play on combinatorial receptive language was greater than the negative effects of seizures by a factor of 17; of sleep problems by a factor of 5; and of high TV exposure by a factor of 3. The positive effect of pretend play on expressive language, sociability, sensory awareness, and health was comparable or smaller than the negative effects of seizures, sleep problems, and high-TV exposure. These results suggest the special role of pretend play in the acquisition of combinatorial receptive language and add to the growing interest in symbolic play interventions (Kasari et al., 2006).

What is the biological reason for such a strong association between combinatorial receptive language and pretend play? One possible explanation is that imaginative play is essential for training neurological networks necessary for acquisition of combinatorial receptive language. Imagination is an umbrella term for multiple neurological mechanisms (Pearson, 2019). Voluntary imagination is always mediated by the lateral prefrontal cortex (LPFC) (Vyshedskiy, 2020) and patients with damage to the LPFC often lose this ability (Dragoy et al., 2017). Conversely, involuntary imagination, such as REM sleep dreaming, does not depend on the LPFC since LPFC is inactive during REM sleep (Braun et al., 1997; Siclari et al., 2017) and patients whose LPFC is damaged do not notice a change in their dreams (Solms, 1997). In addition to completely LPFC-dependent (prefrontal synthesis, integration of modifiers, mental rotation) and unambiguously LPFC-independent mechanisms (REM sleep dreaming, amodal completion), there are hybrid mechanisms as well. For example, the LPFC, known to encode categorical features of objects (Miyashita, 2004; Sidtis et al., 1981), can prime a selected category in memory, without activating any specific memory. Just as in a dream, primed memories can fire randomly and form the basis of *categorically-primed spontaneous imagination*, such as impulsive fantasizing about food or drugs, spontaneous sexual fantasies, racing thoughts about an upcoming exam, an anxiety over a missing child, compulsive jealousy, or spontaneous scientific insights (Vyshedskiy, 2019b).

Voluntary imagination, but not involuntary imagination, is an essential component of language. To understand the difference between “the lion on the tiger” and “the tiger on the lion,” it is not enough to understand the words and the grammar, but it is necessary to *imagine* the lion and the tiger together to appreciate their relations. Linguists refer to this property of human languages as recursion, since it can be used to build nested (recursive) explanations. E.g., a sentence “a snake on the boulder, to the left of the tall tree, that is behind the hill,” forces the interlocutor to use voluntary imagination to combine four objects: a snake, the boulder, the tree, and the hill. Comprehension of spatial prepositions and recursion is impossible without the capacity for voluntary imagination.

Children acquire voluntary imagination and the associated combinatorial recursive language between the age of 3 and 4 years. Based on our research experience, children younger than 3 years did not understand spatial prepositions or recursion, but children older than 4 generally understood both (Vyshedskiy, DuBois, et al., 2020). Pretend play, on the other hand, is initiated between the ages of 1.5 to 2 (Lillard et al., 2011), two years ahead of voluntary imagination acquisition.

The strong association of pretend play and combinatorial receptive language suggests that mechanisms of voluntary imagination take their roots in more primitive mechanisms of pretend play. Neurologically, voluntary imagination is mediated by the LPFC control over the posterior cortex via arcuate fasciculus and other frontoposterior fibers. Voluntary imagination relies on control of timing of neuronal firing in the posterior cortex (Vyshedskiy & Dunn, 2015b). Such precise control of timing is only possible through establishing synchronous frontoposterior networks via finely-tuned myelination of arcuate fasciculus (Vyshedskiy, 2019a, 2019b).

Pretend play falls under the *categorically-primed spontaneous imagination*. Its mechanism is based on the LPFC control over the posterior cortex, but this is a much simpler type of control. In pretend play, the LPFC only primes the categories of objects in memory (Vyshedskiy, 2019b); the activation of mental objects is spontaneous. Posterior cortex neuronal firing timing is *not* controlled. Accordingly, it is possible that pretend play, that occurs universally across all cultures and begins unprompted, can be an essential precursor of voluntary imagination: the simpler *categorically-primed spontaneous imagination* can provide the training necessary for acquisition of more combinatorial voluntary imagination via myelination fine-tuning of arcuate fasciculus and other related mechanisms that aid in establishing a synchronous frontoposterior network.

### MSEC – an important assessment of combinatorial receptive language

Atypically developing children can learn many words without acquiring combinatorial receptive language. An assessment based only on a child’s receptive vocabulary or only on expressive language skills may fail to determine a significant developmental delay in combinatorial receptive language acquisition. In this study, MSEC was significantly more sensitive to effect of pretend play (1.9-fold difference between groups; p <0.0001) compared to the expressive language subscale (1.4-fold difference between groups in subscale 1 of ATEC; p <0.0001) and the sensory awareness subscale (1.3-fold difference between groups in subscale 3 of ATEC; p =0.0009). Additionally, in a 3-year clinical study of 6,454 ASD children, children who engaged with a novel language intervention increased their MSEC score 2.2-fold compared to children with similar initial evaluations (p<0.0001) (Vyshedskiy, DuBois, et al., 2020). At the same time the expressive language score measured by ATEC subscale 1 only increased 1.4-fold and the p-value was significantly lower (0.0144). In another 3-year epidemiological study of the effect of passive video and television watching on 3,227 ASD children, longer video and television watching were associated with 1.3-fold better development of expressive language (p=0.0719) but significantly impeded development of receptive language (1.4-fold difference between the groups; p=0.0128), as measured by MSEC (Fridberg et al., 2021).

This dissociation between receptive and expressive language trajectories observed in all three studies strongly implies the need for a regular assessment of combinatorial receptive language in addition to expressive language. If MSEC evaluation data were not available in the television study, we would conclude that longer video and television watching had a positive effect on ASD children as exhibited by the expressive language subscale. Only addition of the MSEC scale allowed us to detect the significant negative effect of passive video and television watching in ASD children.

### Limitations

Epidemiological monitoring of children provides access to a large number of children at relatively low cost, but has obvious downsides, such as relying on parent reports for diagnosis and assessments. These downsides can be partially mitigated by carefully structured questionnaires administered only a few times a year. We have previously demonstrated that when administered every three months, there was no evaluation bias (Vyshedskiy, Khokhlovich, et al., 2020). While parents may sometimes yield to wishful thinking and overestimate their children’s abilities on a single assessment (Scattone et al., 2011), the pattern of changes generated by measuring the score dynamics over multiple assessments, provide meaningful data on a child’s developmental trajectory.

Another limitation is that this study observational design cannot definitively prove causality since not all confounders can be adjusted appropriately. Our results only inform on the strong association of pretend play with combinatorial language. Future randomized controlled studies of pretend play interventions will be able to answer the question of causality between pretend play and a child’s developmental trajectory.

## Data Availability

All data produced in the present study are available upon reasonable request to the authors

## Funding

This research received no external funding.

## Acknowledgements

We wish to thank Dr. Petr Ilyinskii for his scrupulous editing of this manuscript.

## Author contributions

AV and EK designed the study. AV analyzed the data. AV wrote the paper.

## Competing Interests

Authors declare no competing interests.

## Informed Consent

Caregivers have consented to anonymized data analysis and publication of the results. The study was conducted in compliance with the Declaration of Helsinki (Association, 2013).

## Compliance with Ethical Standards

Using the Department of Health and Human Services regulations found at 45 CFR 46.101(b)(4), the Biomedical Research Alliance of New York LLC Institutional Review Board (IRB) determined that this research project is exempt from IRB oversight.

## Data Availability

De-identified raw data from this manuscript are available from the corresponding author upon reasonable request.

## Code availability statement

Code is available from the corresponding author upon reasonable request.

## Notes

### Competing Interest Statement

The authors have declared no competing interest.

### Funding Statement

This study did not receive any funding

## References

Al Backer, N. B. (2016). Correlation between Autism Treatment Evaluation Checklist (ATEC) and Childhood Autism Rating Scale (CARS) in the evaluation of autism spectrum disorder. Sudanese Journal of Paediatrics, 16(1), 17–22.

Association, W. M. (2013). World Medical Association Declaration of Helsinki: Ethical principles for medical research involving human subjects. Jama, 310(20), 2191–2194.

Baron-Cohen, S., Allen, J., & Gillberg, C. (1992). Can autism be detected at 18 months?: The needle, the haystack, and the CHAT. The British Journal of Psychiatry, 161(6), 839–843.

Braun, A. R., Balkin, T. J., Wesensten, N. J., Carson, R. E., Varga, M., Baldwin, P., Selbie, S., Belenky, G., & Herscovitch, P. (1997). Regional cerebral blood flow throughout the sleep-wake cycle. Brain, 120(7), 1173–1197.

Braverman, J., Dunn, R., & Vyshedskiy, A. (2018). Development of the Mental Synthesis Evaluation Checklist (MSEC): A Parent-Report Tool for Mental Synthesis Ability Assessment in Children with Language Delay. Children, 5(5), 62. https://doi.org/10.3390/children5050062

Charman, T., Howlin, P., Berry, B., & Prince, E. (2004). Measuring developmental progress of children with autism spectrum disorder on school entry using parent report. Autism, 8(1), 89–100.

Dragoy, O., Akinina, Y., & Dronkers, N. (2017). Toward a functional neuroanatomy of semantic aphasia: A history and ten new cases. Cortex, 97, 164–182.

Dunn, R., Elgart, J., Lokshina, L., Faisman, A., Khokhlovich, E., Gankin, Y., & Vyshedskiy, A. (2017a). Children With Autism Appear To Benefit From Parent-Administered Computerized Cognitive And Language Exercises Independent Of the Child’s Age Or Autism Severity. Autism Open Access, 7(217). https://doi.org/10.4172/2165-7890.1000217

Dunn, R., Elgart, J., Lokshina, L., Faisman, A., Khokhlovich, E., Gankin, Y., & Vyshedskiy, A. (2017b). Comparison of performance on verbal and nonverbal multiple-cue responding tasks in children with ASD. Autism Open Access, 7, 218. https://doi.org/10.4172/2165-7890.1000218

Dunn, R., Elgart, J., Lokshina, L., Faisman, A., Waslick, M., Gankin, Y., & Vyshedskiy, A. (2017). Tablet-Based Cognitive Exercises as an Early Parent-Administered Intervention Tool for Toddlers with Autism—Evidence from a Field Study. Clinical Psychiatry, 3(1). https://doi.org/10.21767/2471-9854.100037

Forman, P., Edward, E., & Vyshedskiy, A. (2022). Effect of seizures on developmental trajectories in children with autism. MedRxiv, 2022.03.30.22273179. https://doi.org/10.1101/2022.03.30.22273179

Fridberg, E., Khokhlovich, E., & Vyshedskiy, A. (2021). Watching Videos and Television Is Related to a Lower Development of Complex Language Comprehension in Young Children with Autism. Healthcare, 9(4), 423.

Geier, D. A., Kern, J. K., & Geier, M. R. (2013). A comparison of the Autism Treatment Evaluation Checklist (ATEC) and the Childhood Autism Rating Scale (CARS) for the quantitative evaluation of autism. Journal of Mental Health Research in Intellectual Disabilities, 6(4), 255–267.

Jagadeesan, P., Kabbani, P., & Vyshedskiy, A. (n.d.). Parent-reported assessment scores reflect ASD severity in 2-to 7-year-old children. Under Review.

Jarusiewicz, B. (2002). Efficacy of Neurofeedback for Children in the Autistic Spectrum: A Pilot Study. Journal of Neurotherapy, 6(4), 39–49. https://doi.org/10.1300/J184v06n04_05

Kasari, C., Freeman, S., & Paparella, T. (2006). Joint attention and symbolic play in young children with autism: A randomized controlled intervention study. Journal of Child Psychology and Psychiatry, 47(6), 611–620.

Klaveness, J., Bigam, J., & Reichelt, K. L. (2013). The varied rate of response to dietary intervention in autistic children. Open Journal of Psychiatry, 3(02), 56.

Levin, J., Khokhlovich, E., & Vyshedskiy, A. (2022). Sleep problems effect on developmental trajectories in children with autism. MedRxiv, 2022.03.30.22273178. https://doi.org/10.1101/2022.03.30.22273178

Lewis, V. (2000). Relationships between symbolic play, functional play, verbal and non-verbal ability in young children. International Journal of Language & Communication Disorders, 35(1), 117–127.

Lillard, A. S., Pinkham, A. M., & Smith, E. (2011). Pretend play and cognitive development. The Wiley-Blackwell Handbook of Childhood Cognitive Development, 32, 285.

Lord, C., Risi, S., Lambrecht, L., Cook, E. H., Leventhal, B. L., DiLavore, P. C., Pickles, A., & Rutter, M. (2000). The Autism Diagnostic Observation Schedule—Generic: A standard measure of social and communication deficits associated with the spectrum of autism. Journal of Autism and Developmental Disorders, 30(3), 205–223.

Magiati, I., Moss, J., Yates, R., Charman, T., & Howlin, P. (2011). Is the Autism Treatment Evaluation Checklist a useful tool for monitoring progress in children with autism spectrum disorders? Journal of Intellectual Disability Research, 55(3), 302–312. https://doi.org/10.1111/j.1365-2788.2010.01359.x

Mahapatra, S., Khokhlovich, E., Martinez, S., Kannel, B., Edelson, S. M., & Vyshedskiy, A. (2018). Longitudinal Epidemiological Study of Autism Subgroups Using Autism Treatment Evaluation Checklist (ATEC) Score. Autism and Developmental Disorders, 1(12). https://doi.org/10.1007/s10803-018-3699-2

Mahapatra, S., Vyshedsky, D., Martinez, S., Kannel, B., Braverman, J., Edelson, S. M., & Vyshedskiy, A. (2018). Autism Treatment Evaluation Checklist (ATEC) norms: A “growth chart” for ATEC score changes as a function of age. Children, 5(2). https://doi.org/10.3390

Miyashita, Y. (2004). Cognitive memory: Cellular and network machineries and their top-down control. Science, 306(5695), 435–440.

Osterling, J., & Dawson, G. (1994). Early recognition of children with autism: A study of first birthday home videotapes. Journal of Autism and Developmental Disorders, 24(3), 247–257.

Pearson, J. (2019). The human imagination: The cognitive neuroscience of visual mental imagery. Nature Reviews Neuroscience, 624–634.

Rimland, B., & Edelson, S. (1999). Autism Research Institute. Autism Treatment Evaluation Checklist (ATEC).

Scattone, D., Raggio, D. J., & May, W. (2011). Comparison of the vineland adaptive behavior scales, and the bayley scales of infant and toddler development. Psychological Reports, 109(2), 626–634.

Schneider, B., Carnoy, M., Kilpatrick, J., Schmidt, W. H., & Shavelson, R. J. (2007). Estimating causal effects using experimental and observational design. American Educational & Reseach Association.

Siclari, F., Baird, B., Perogamvros, L., Bernardi, G., LaRocque, J. J., Riedner, B., Boly, M., Postle, B. R., & Tononi, G. (2017). The neural correlates of dreaming. Nature Neuroscience, 20(6), 872.

Sidtis, J. J., Volpe, B. T., Holtzman, J. D., Wilson, D. H., & Gazzaniga, M. S. (1981). Cognitive interaction after staged callosal section: Evidence for transfer of semantic activation. Science, 212(4492), 344–346.

Solms, M. (1997). The neuropsychology of dreams: A clinico-anatomical study. L. Erlbaum. http://www.cell.com/trends/cognitive-sciences/pdf/S1364-6613(98)01166-8.pdf

Stagnitti, K., & Lewis, F. M. (2015). Quality of pre-school children’s pretend play and subsequent development of semantic organization and narrative re-telling skills. International Journal of Speech-Language Pathology, 17(2), 148–158.

Toth, K., Munson, J., Meltzoff, A. N., & Dawson, G. (2006). Early predictors of communication development in young children with autism spectrum disorder: Joint attention, imitation, and toy play. Journal of Autism and Developmental Disorders, 36(8), 993–1005.

Vyshedskiy. (2019a). Language evolution to revolution: The leap from rich-vocabulary non-recursive communication system to recursive language 70,000 years ago was associated with acquisition of a novel component of imagination, called Prefrontal Synthesis, enabled by a mutation that slowed down the prefrontal cortex maturation simultaneously in two or more children – the Romulus and Remus hypothesis. Research Ideas and Outcomes, 5, e38546. https://doi.org/10.3897/rio.5.e38546

Vyshedskiy, A. (2019b). gNeuroscience of imagination and implications for hominin evolution. Journal of Current Neurobiology, 10(2), 89–109. https://doi.org/10.31234/osf.io/skxwc

Vyshedskiy, A. (2020). Voluntary and Involuntary Imagination: Neurological Mechanisms, Developmental Path, Clinical Implications, and Evolutionary Trajectory. Evolutionary Studies in Imaginative Culture, 4(2), 1–17. https://doi.org/10.26613/esic/4.2.186

Vyshedskiy, A., DuBois, M., Mugford, E., Piryatinsky, I., Radi, K., Braverman, J., & Maslova, V. (2020). Novel Linguistic Evaluation of Prefrontal Synthesis (LEPS) test measures prefrontal synthesis acquisition in neurotypical children and predicts high-functioning versus low-functioning class assignment in individuals with autism. Applied Neuropsychology: Child. https://doi.org/10.1080/21622965.2020.1758700

Vyshedskiy, A., & Dunn, R. (2015). Mental Imagery Therapy for Autism (MITA)-An Early Intervention Computerized Brain Training Program for Children with ASD. Autism Open Access, 5(1000153), 2.

Vyshedskiy, A., Khokhlovich, E., Dunn, R., Faisman, A., Elgart, J., Lokshina, L., Gankin, Y., Ostrovsky, S., deTorres, L., & Edelson, S. M. (2020). Novel prefrontal synthesis intervention improves language in children with autism. Healthcare, 8(4), 566. https://doi.org/doi.org/10.3390/healthcare8040566

Vyshedskiy, & Dunn, R. (2015). Mental synthesis involves the synchronization of independent neuronal ensembles. Research Ideas and Outcomes, 1, e7642.

Whitehouse, A. J., Granich, J., Alvares, G., Busacca, M., Cooper, M. N., Dass, A., Duong, T., Harper, R., Marshall, W., Richdale, A., & others. (2017). A randomised controlled trial of an iPad-based application to complement early behavioural intervention in Autism Spectrum Disorder. Journal of Child Psychology and Psychiatry, 58(9), 1042–1052.

