## Supplementary Material for "Pretend play predicts receptive and expressive language trajectories in young children with autism"

**Table S1: LS Means (SE; 95% CI) for Receptive Language MSEC subscale score. The difference between Pretend Play and no Pretend Play (PP – noPP) is presented as: LS Mean (SE; P-value). The negative PP – noPP difference indicates that the noPP group had higher score and therefore more severe symptoms.**

| Visit Number | Pretend Play group | No Pretend Play group | PP – noPP |
|---|---|---|---|
| Baseline | 27 (0.134; 26.7 - 27.2) | 28.8 (0.13; 28.5 - 29.1) | -1.83 (0.18; <0.0001) |
| Month 6 | 24.8 (0.16; 24.5 - 25.2) | 28.5 (0.156; 28.2 - 28.9) | -3.7 (0.22; <0.0001) |
| Month 9 | 23.5 (0.163; 23.2 - 23.8) | 27.8 (0.159; 27.5 - 28.1) | -4.32 (0.22; <0.0001) |
| Month 12 | 23.3 (0.181; 22.9 - 23.6) | 27 (0.182; 26.7 - 27.4) | -3.75 (0.25; <0.0001) |
| Month 15 | 21.7 (0.217; 21.3 - 22.1) | 26.4 (0.211; 26 - 26.9) | -4.77 (0.3; <0.0001) |
| Month 18 | 21.2 (0.261; 20.7 - 21.7) | 26.3 (0.251; 25.8 - 26.8) | -5.06 (0.36; <0.0001) |
| Month 21 | 20 (0.304; 19.4 - 20.6) | 25.2 (0.275; 24.7 - 25.8) | -5.22 (0.41; <0.0001) |
| Month 24 | 19.5 (0.328; 18.9 - 20.2) | 25.3 (0.304; 24.7 - 25.9) | -5.77 (0.45; <0.0001) |
| Month 27 | 18.5 (0.363; 17.8 - 19.2) | 24.8 (0.335; 24.1 - 25.5) | -6.29 (0.49; <0.0001) |
| Month 30 | 17.2 (0.416; 16.4 - 18) | 24.3 (0.361; 23.6 - 25) | -7.1 (0.55; <0.0001) |
| Month 33 | 16.9 (0.477; 16 - 17.9) | 24.2 (0.42; 23.4 - 25) | -7.25 (0.63; <0.0001) |
| Month 36 | 15.7 (0.546; 14.6 - 16.7) | 22.9 (0.474; 22 - 23.8) | -7.26 (0.72; <0.0001) |
| Month 36 - Baseline | -11.31 (0.55; <0.0001) | -5.88 (0.48; <0.0001) | na |

**Table S2: LS Means (SE; 95% CI) for Expressive Language measured by the Subscale 1 of ATEC. The difference between Pretend Play and no Pretend Play (PP – noPP) is presented as: LS Mean (SE; P-value). The negative PP – noPP difference indicates that the noPP group had higher score and therefore more severe symptoms.**

| Visit Number | Pretend Play group | No Pretend Play group | PP – noPP |
|---|---|---|---|
| Baseline | 14.98 (0.0993; 14.78 - 15.17) | 15.77 (0.0971; 15.58 - 15.96) | -0.79 (0.13; <0.0001) |
| Month 6 | 12.59 (0.1182; 12.36 - 12.82) | 14.59 (0.1165; 14.36 - 14.82) | -2 (0.16; <0.0001) |
| Month 9 | 11.59 (0.1202; 11.35 - 11.82) | 13.76 (0.1184; 13.53 - 13.99) | -2.17 (0.16; <0.0001) |
| Month 12 | 10.86 (0.1339; 10.59 - 11.12) | 12.94 (0.135; 12.67 - 13.2) | -2.08 (0.19; <0.0001) |
| Month 15 | 9.78 (0.1605; 9.47 - 10.1) | 12.18 (0.1562; 11.87 - 12.48) | -2.39 (0.22; <0.0001) |
| Month 18 | 9.26 (0.1925; 8.88 - 9.63) | 12.1 (0.1857; 11.73 - 12.46) | -2.84 (0.26; <0.0001) |
| Month 21 | 8.73 (0.224; 8.29 - 9.17) | 11.21 (0.2028; 10.81 - 11.61) | -2.48 (0.3; <0.0001) |
| Month 24 | 8.29 (0.242; 7.81 - 8.76) | 10.91 (0.2244; 10.47 - 11.35) | -2.63 (0.33; <0.0001) |
| Month 27 | 8.11 (0.2675; 7.59 - 8.64) | 11.04 (0.2468; 10.56 - 11.53) | -2.93 (0.36; <0.0001) |
| Month 30 | 7.22 (0.3064; 6.62 - 7.82) | 10.69 (0.2656; 10.17 - 11.21) | -3.47 (0.4; <0.0001) |
| Month 33 | 7.69 (0.3506; 7 - 8.37) | 10.39 (0.309; 9.79 - 11) | -2.71 (0.47; <0.0001) |
| Month 36 | 7.3 (0.4018; 6.51 - 8.09) | 10.25 (0.3488; 9.57 - 10.94) | -2.95 (0.53; <0.0001) |
| Month 36 - Baseline | -7.68 (0.4; <0.0001) | -5.52 (0.35; <0.0001) | na |

**Table S3: LS Means (SE; 95% CI) for Sociability subscale score measured by the Subscale 2 of ATEC. The difference between Pretend Play and no Pretend Play (PP – noPP) is presented as: LS Mean (SE; P-value). The negative PP – noPP difference indicates that the noPP group had higher score and therefore more severe symptoms.**

| Visit Number | Pretend Play group | No Pretend Play group | PP – noPP |
|---|---|---|---|
| Baseline | 13.5 (0.127; 13.2 - 13.7) | 13.4 (0.124; 13.1 - 13.6) | 0.13 (0.17; 0.454) |
| Month 6 | 12.6 (0.154; 12.3 - 12.9) | 13.1 (0.151; 12.8 - 13.4) | -0.56 (0.21; 0.0068) |
| Month 9 | 12.3 (0.156; 12 - 12.6) | 13 (0.154; 12.7 - 13.3) | -0.65 (0.21; 0.0021) |
| Month 12 | 11.7 (0.176; 11.3 - 12) | 12.6 (0.177; 12.2 - 12.9) | -0.92 (0.24; 0.0001) |
| Month 15 | 11.9 (0.212; 11.5 - 12.4) | 12.3 (0.206; 11.9 - 12.7) | -0.37 (0.29; 0.2009) |
| Month 18 | 11.3 (0.256; 10.8 - 11.8) | 12.5 (0.247; 12 - 13) | -1.21 (0.35; 0.0006) |
| Month 21 | 11.1 (0.299; 10.5 - 11.7) | 12.4 (0.27; 11.9 - 12.9) | -1.29 (0.4; 0.0013) |
| Month 24 | 11.6 (0.324; 11 - 12.3) | 12.1 (0.3; 11.5 - 12.7) | -0.46 (0.44; 0.2917) |
| Month 27 | 11.4 (0.359; 10.7 - 12.1) | 12.4 (0.331; 11.7 - 13) | -0.95 (0.49; 0.0518) |
| Month 30 | 11.1 (0.412; 10.3 - 11.9) | 12 (0.357; 11.3 - 12.7) | -0.9 (0.54; 0.098) |
| Month 33 | 11.3 (0.472; 10.3 - 12.2) | 12.5 (0.416; 11.7 - 13.3) | -1.2 (0.63; 0.0554) |
| Month 36 | 10.9 (0.541; 9.8 - 11.9) | 11.1 (0.47; 10.2 - 12) | -0.24 (0.72; 0.7348) |
| Month 36 - Baseline | -2.63 (0.55; <0.0001) | -2.26 (0.48; <0.0001) | na |

**Table S4: LS Means (SE; 95% CI) for the Sensory/Cognitive Awareness subscale score measured by the Subscale 3 of ATEC. The difference between Pretend Play and no Pretend Play (PP – noPP) is presented as: LS Mean (SE; P-value). The negative PP – noPP difference indicates that the noPP group had higher score and therefore more severe symptoms.**

| Visit Number | Pretend Play group | No Pretend Play group | PP – noPP |
|---|---|---|---|
| Baseline | 12.77 (0.104; 12.56 - 12.97) | 14.15 (0.103; 13.95 - 14.35) | -1.39 (0.14; <0.0001) |
| Month 6 | 11.89 (0.126; 11.65 - 12.14) | 14.76 (0.126; 14.51 - 15.01) | -2.87 (0.17; <0.0001) |
| Month 9 | 11.49 (0.129; 11.23 - 11.74) | 14.49 (0.128; 14.24 - 14.74) | -3.01 (0.18; <0.0001) |
| Month 12 | 11.2 (0.145; 10.92 - 11.49) | 14.14 (0.147; 13.85 - 14.42) | -2.93 (0.2; <0.0001) |
| Month 15 | 10.95 (0.176; 10.6 - 11.29) | 13.65 (0.172; 13.31 - 13.99) | -2.7 (0.24; <0.0001) |
| Month 18 | 10.25 (0.213; 9.83 - 10.67) | 14.03 (0.206; 13.63 - 14.43) | -3.78 (0.29; <0.0001) |
| Month 21 | 10.29 (0.25; 9.8 - 10.78) | 13.41 (0.226; 12.97 - 13.85) | -3.12 (0.33; <0.0001) |
| Month 24 | 9.96 (0.27; 9.43 - 10.49) | 13.49 (0.251; 13 - 13.98) | -3.53 (0.37; <0.0001) |
| Month 27 | 9.2 (0.3; 8.61 - 9.78) | 13.74 (0.277; 13.2 - 14.28) | -4.54 (0.41; <0.0001) |
| Month 30 | 9.43 (0.345; 8.75 - 10.11) | 13.09 (0.298; 12.51 - 13.68) | -3.66 (0.45; <0.0001) |
| Month 33 | 8.94 (0.395; 8.16 - 9.71) | 12.49 (0.348; 11.81 - 13.17) | -3.55 (0.53; <0.0001) |
| Month 36 | 10.35 (0.453; 9.46 - 11.24) | 12.34 (0.393; 11.57 - 13.11) | -1.99 (0.6; 0.0009) |
| Month 36 - Baseline | -2.41 (0.46; <0.0001) | -1.81 (0.4; <0.0001) | na |

**Table S5: LS Means (SE; 95% CI) for Health/Physical/Behavior subscale score measured by the Subscale 4 of ATEC. The difference between Pretend Play and no Pretend Play (PP – noPP) is presented as: LS Mean (SE; P-value). The negative PP – noPP difference indicates that the noPP group had higher score and therefore more severe symptoms.**

| Visit Number | Pretend Play group | No Pretend Play group | PP – noPP |
|---|---|---|---|
| Baseline | 21.4 (0.193; 21 - 21.8) | 20.3 (0.187; 19.9 - 20.6) | 1.11 (0.25; <0.0001) |
| Month 6 | 20.6 (0.232; 20.2 - 21.1) | 20 (0.227; 19.6 - 20.5) | 0.57 (0.31; 0.0695) |
| Month 9 | 20.7 (0.236; 20.3 - 21.2) | 20.2 (0.232; 19.7 - 20.6) | 0.54 (0.32; 0.0887) |
| Month 12 | 20.2 (0.265; 19.7 - 20.7) | 20.5 (0.266; 20 - 21) | -0.3 (0.37; 0.4144) |
| Month 15 | 20.3 (0.32; 19.7 - 20.9) | 19.1 (0.31; 18.5 - 19.7) | 1.17 (0.44; 0.0075) |
| Month 18 | 19.4 (0.386; 18.7 - 20.2) | 19.7 (0.371; 19 - 20.5) | -0.3 (0.53; 0.5655) |
| Month 21 | 19 (0.451; 18.1 - 19.9) | 19.6 (0.407; 18.8 - 20.4) | -0.63 (0.6; 0.295) |
| Month 24 | 20.3 (0.488; 19.3 - 21.2) | 19 (0.451; 18.1 - 19.9) | 1.26 (0.66; 0.056) |
| Month 27 | 19.2 (0.54; 18.2 - 20.3) | 19.6 (0.497; 18.6 - 20.6) | -0.37 (0.73; 0.615) |
| Month 30 | 18.9 (0.621; 17.7 - 20.2) | 18.4 (0.536; 17.4 - 19.5) | 0.53 (0.82; 0.5188) |
| Month 33 | 19.2 (0.711; 17.9 - 20.6) | 19.2 (0.625; 18 - 20.4) | 0.06 (0.94; 0.9533) |
| Month 36 | 20.0 (0.815; 18.5 - 21.6) | 19.3 (0.707; 17.9 - 20.7) | 0.76 (1.08; 0.4782) |
| Month 36 - Baseline | -1.33 (0.82; 0.1062) | -0.98 (0.72; 0.1699) | na |

**Table S6: LS Means (SE; 95% CI) for Receptive Language MSEC subscale score when controlling for expressive language. The difference between Pretend Play and no Pretend Play (PP – noPP) is presented as: LS Mean (SE; P-value). The negative PP – noPP difference indicates that the noPP group had higher score and therefore more severe symptoms.**

| Visit Number | Pretend Play group | No Pretend Play group | PP – noPP |
|---|---|---|---|
| Baseline | 25.9 (0.126; 25.7 - 26.2) | 27.3 (0.124; 27 - 27.5) | -1.36 (0.17; <0.0001) |
| Month 6 | 24.7 (0.149; 24.4 - 25) | 27.5 (0.148; 27.2 - 27.7) | -2.77 (0.2; <0.0001) |
| Month 9 | 23.7 (0.152; 23.4 - 24) | 27.1 (0.15; 26.8 - 27.3) | -3.32 (0.21; <0.0001) |
| Month 12 | 23.8 (0.17; 23.5 - 24.1) | 26.6 (0.171; 26.2 - 26.9) | -2.78 (0.24; <0.0001) |
| Month 15 | 22.6 (0.206; 22.2 - 23) | 26.3 (0.199; 25.9 - 26.7) | -3.67 (0.28; <0.0001) |
| Month 18 | 22.3 (0.248; 21.9 - 22.8) | 26.2 (0.238; 25.7 - 26.7) | -3.85 (0.34; <0.0001) |
| Month 21 | 21.3 (0.29; 20.7 - 21.9) | 25.5 (0.261; 25 - 26) | -4.18 (0.39; <0.0001) |
| Month 24 | 21 (0.314; 20.4 - 21.7) | 25.6 (0.289; 25.1 - 26.2) | -4.6 (0.42; <0.0001) |
| Month 27 | 20.1 (0.347; 19.4 - 20.8) | 25.1 (0.319; 24.5 - 25.7) | -5.02 (0.47; <0.0001) |
| Month 30 | 19 (0.399; 18.3 - 19.8) | 24.8 (0.344; 24.1 - 25.5) | -5.76 (0.52; <0.0001) |
| Month 33 | 18.6 (0.456; 17.7 - 19.5) | 24.8 (0.401; 24 - 25.6) | -6.24 (0.61; <0.0001) |
| Month 36 | 17.5 (0.523; 16.5 - 18.5) | 23.6 (0.453; 22.7 - 24.5) | -6.09 (0.69; <0.0001) |
| Month 36 - Baseline | -8.42 (0.53; <0.0001) | -3.69 (0.46; <0.0001) | na |

**Table S7: LS Means (SE; 95% CI) for Expressive Language score when controlling for receptive language. The difference between Pretend Play and no Pretend Play (PP – noPP) is presented as: LS Mean (SE; P-value). The negative PP – noPP difference indicates that the noPP group had higher score and therefore more severe symptoms.**

| Visit Number | Pretend Play group | No Pretend Play group | PP – noPP |
|---|---|---|---|
| Baseline | 14.86 (0.0943; 14.67 - 15.04) | 14.99 (0.0943; 14.81 - 15.18) | -0.13 (0.13; 0.2911) |
| Month 6 | 12.9 (0.1128; 12.67 - 13.12) | 13.86 (0.1125; 13.64 - 14.09) | -0.97 (0.16; <0.0001) |
| Month 9 | 12.17 (0.1154; 11.95 - 12.4) | 13.18 (0.1138; 12.96 - 13.4) | -1.01 (0.16; <0.0001) |
| Month 12 | 11.48 (0.1288; 11.23 - 11.74) | 12.51 (0.1294; 12.25 - 12.76) | -1.02 (0.18; <0.0001) |
| Month 15 | 10.7 (0.1552; 10.4 - 11.01) | 11.87 (0.1496; 11.58 - 12.16) | -1.17 (0.21; <0.0001) |
| Month 18 | 10.24 (0.186; 9.87 - 10.6) | 11.8 (0.1781; 11.45 - 12.15) | -1.56 (0.26; <0.0001) |
| Month 21 | 9.97 (0.217; 9.55 - 10.4) | 11.13 (0.1943; 10.75 - 11.51) | -1.16 (0.29; <0.0001) |
| Month 24 | 9.63 (0.2346; 9.17 - 10.09) | 10.85 (0.2152; 10.43 - 11.27) | -1.23 (0.32; 0.0001) |
| Month 27 | 9.65 (0.2596; 9.14 - 10.16) | 11.07 (0.2368; 10.61 - 11.53) | -1.42 (0.35; <0.0001) |
| Month 30 | 9.06 (0.2978; 8.48 - 9.65) | 10.8 (0.2549; 10.3 - 11.3) | -1.73 (0.39; <0.0001) |
| Month 33 | 9.59 (0.3401; 8.93 - 10.26) | 10.5 (0.2967; 9.92 - 11.09) | -0.91 (0.45; 0.0432) |
| Month 36 | 9.47 (0.3898; 8.7 - 10.23) | 10.61 (0.3351; 9.96 - 11.27) | -1.15 (0.51; 0.0253) |
| Month 36 - Baseline | -5.39 (0.39; <0.0001) | -4.38 (0.34; <0.0001) | na |
